# Real-time Nowcasting and Forecasting of COVID-19 Dynamics in England: the first wave?

**DOI:** 10.1101/2020.08.24.20180737

**Authors:** Paul Birrell, Joshua Blake, Edwin van Leeuwen, PHE Joint Modelling Cell, Nick Gent, Daniela De Angelis

## Abstract

England has been heavily affected by the SARS-CoV-2 pandemic, with severe ‘lock-down’ mitigation measures now gradually being lifted. The real-time pandemic monitoring presented here has contributed to the evidence informing this pandemic management. Estimates on the 10th May showed lock-down had reduced transmission by 75%, the reproduction number falling from 2.6 to 0.61. This regionally-varying impact was largest in London of 81% (95% CrI: 77%–84%). Reproduction numbers have since slowly increased, and on 19th June the probability that the epidemic is growing was greater than 5% in two regions, South West and London. An estimated 8% of the population had been infected, with a higher proportion in London (17%). The infection-to-fatality ratio is 1.1% (0.9%–1.4%) overall but 17% (14%–22%) among the over-75s. This ongoing work will be key to quantifying any widespread resurgence should accrued immunity and effective contact tracing be insufficient to preclude a second wave.

## 1 Introduction

Since the 31st of December 2019, when the government of Wuhan reported treating cases of pneumonia of unknown cause (*1*), more than 13 million individuals have been reported as being infected by SARS-COV-2 globally and over 500,000 have died (*2*). Spread of SARS-COV-2 from Wuhan to other Chinese provinces, Thailand, Japan and the Republic of Korea rapidly occurred in the first 3 weeks of January 2020 (*3*), with an early incursion into Europe centred around clusters in Bavaria, Germany and Haute-Savoie, France, both linked to subsequent cases in Spain (*4*). This international spread eventually led the World Health Organisation to declare a pandemic on 11^th^ March, 2020 (*5*). In the UK, pandemic preparedness plans, developed since 2009 A/H1N1pdm, were rapidly activated with governmental emergency bodies and advisory groups convening before the end of January (*6*).

The UK Response to the COVID-19 pandemic escalated from an initial containment effort to the suppression or ‘lock-down’ strategy introduced on the 23^rd^ of March (*7*). Over this period, through participation in governmental advisory groups, scientists from a number of research institutions fed into the pandemic decision-making processes. Quantitative understanding of the pandemic was based on the work of SPI-M (Scientific Pandemic Influenza Sub-Group on Modelling) (*8*). This work has informed the various phases of the UK response, from constructing planning scenarios for the health system, to monitoring these scenarios through regular now-casting of new infections and forecasting of severe disease and health service demand, see (*9,10*) and papers available from (*8*).

Here we report the work of one of the participating groups, the Public Health England (PHE)/University of Cambridge modelling group. This collaboration, funded to develop modelling methodology for real-time pandemic influenza monitoring (*11*), was re-activated for the COVID-19 pandemic (*12*). The age and spatially structured transmission model developed for influenza (*13,14*), was re-purposed and, as data have increasingly become available, continuously adapted to SARS-COV-2 epidemiology, see Supplementary Materials (SM) ‘Transmission Model’.

The model is implemented through a Bayesian statistical analysis of pandemic surveillance data, incorporating information on the natural history of infection from emerging literature. The surveillance data used are age- and region-specific counts of deaths of people with a lab-confirmed CoVID-19 diagnosis (see Fig. 1A-B); and, from 21^st^ April onwards, weekly batches of serological data, indicating the fraction of the population carrying CoVID-19 antibodies, from NHS Blood and Transplant (NHSBT) samples (see Fig. 1C) (*15*). The Wuhan outbreak additionally informs estimation of model parameters: the duration of infectiousness, the mean time from infection to symptom onset (*16*); and mean time from symptoms onset to death (*17*). Central to age-specific epidemic modelling, contact patterns between age groups have been derived from the POLYMOD study (*18*) stratified by setting (school, workplace, leisure etc), with these matrices sequentially updated using the Google mobility study and the UK time-use survey (*19*), to quantify the change in population mobility and access to these contact settings over time.

**Figure 1:**
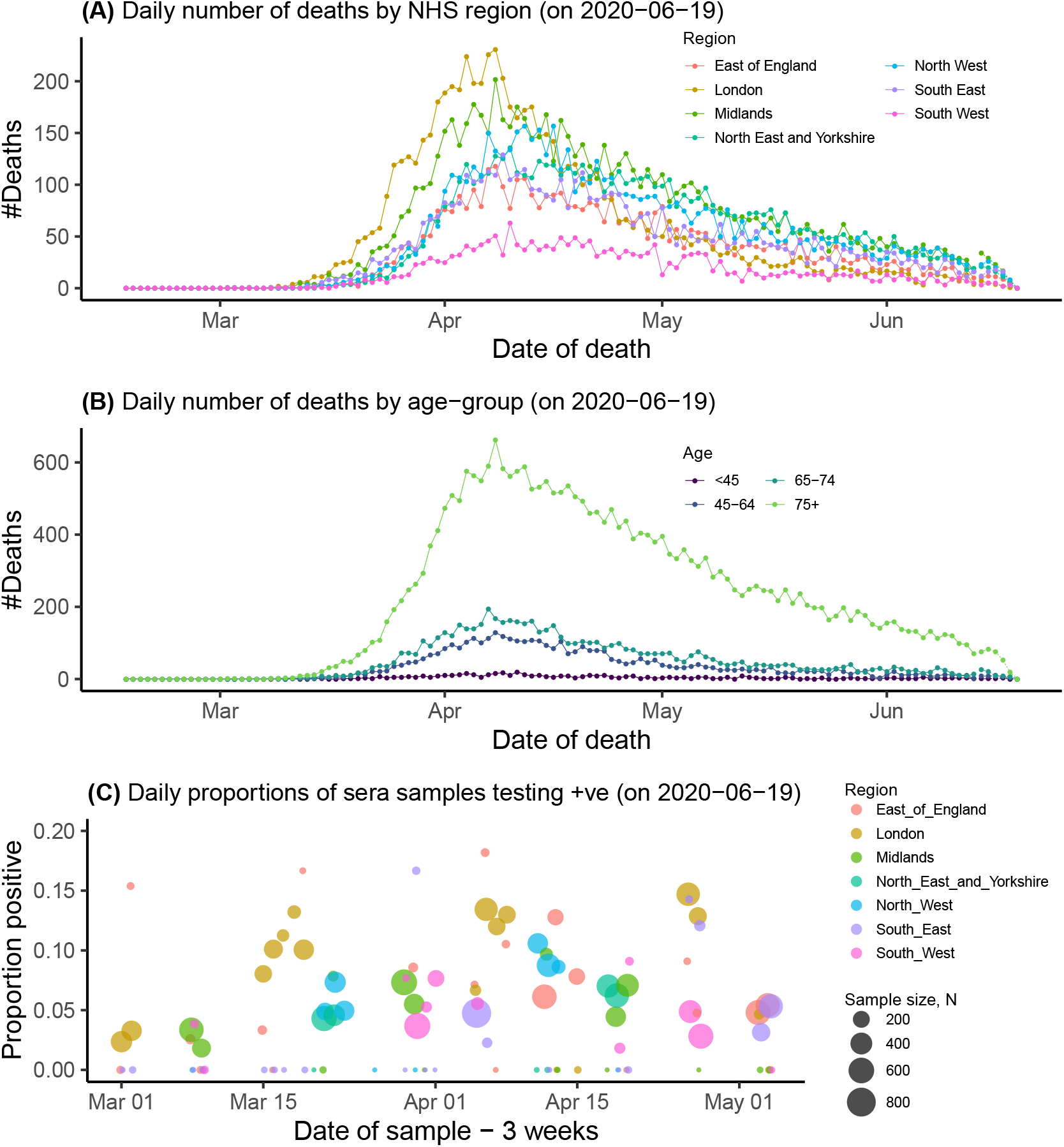
(A) Data on deaths by region, and (B) age; (C) Serological positivity by region and sampling date.

This transmission model has been used consistently throughout the pandemic. Through this regular monitoring we have been able to: anticipate and understand the impact of the lock-down; provide sequential updates of the pandemic dynamics, by estimating the basic (*R*_0_) and effective (*R_t_*) reproduction numbers, (*i.e*. the average number of individuals infected by an infectious individual in: a totally susceptible population with no control measures; and a partially susceptible population under some control measures respectively); and inform the gradual relaxation of the lock-down.

Here we focus on the contribution of our work at specific dates spanning three key periods: pre-lock-down (before 23^rd^ of March), lock-down (until 11^th^ May); and two subsequent dates that allow the assessment of the gradual easing of the lockdown.

### Pre-lock-down

After initial attempts to contain the pandemic through trace and test strategies (*20*) and to mitigate the burden on the National Health Service (NHS) through combinations of non-pharmaceutical interventions (*e.g*. case isolation, restrictions on foreign travel, shielding of vulnerable groups, cancellation of mass gatherings), the pressing question became: what level of stringent social distancing measures would be necessary to suppress transmission? At this stage, infection was not sufficiently widespread in each of the seven NHS regions for the data to inform a fully stratified model, by this time there had only been four deaths in the whole of the South-West. Therefore, no age structure was used and the country was stratified into two regions: London, where the number of deaths was significantly higher (Fig 1A), and Outside London. Assuming a pandemic intervention is imposed on 23^rd^ March, the model was fitted to data on CoVID-19 confirmed deaths to 15^th^ March, and then used to project epidemic curves forward a further eight weeks. These projections assumed differing reductions in contact rates (24%, 48%, 64%) (see Equation (*6*), SM) and consequently transmission.

Fig. 2 shows the projections for different levels of this reduction. The dashed red vertical line shows the date of the most recent data included in the analysis and the dashed purple line represents the timing of the intervention. Each column shows the projected epidemic infection and death rates under three assumed intervention impacts.

**Figure 2:**
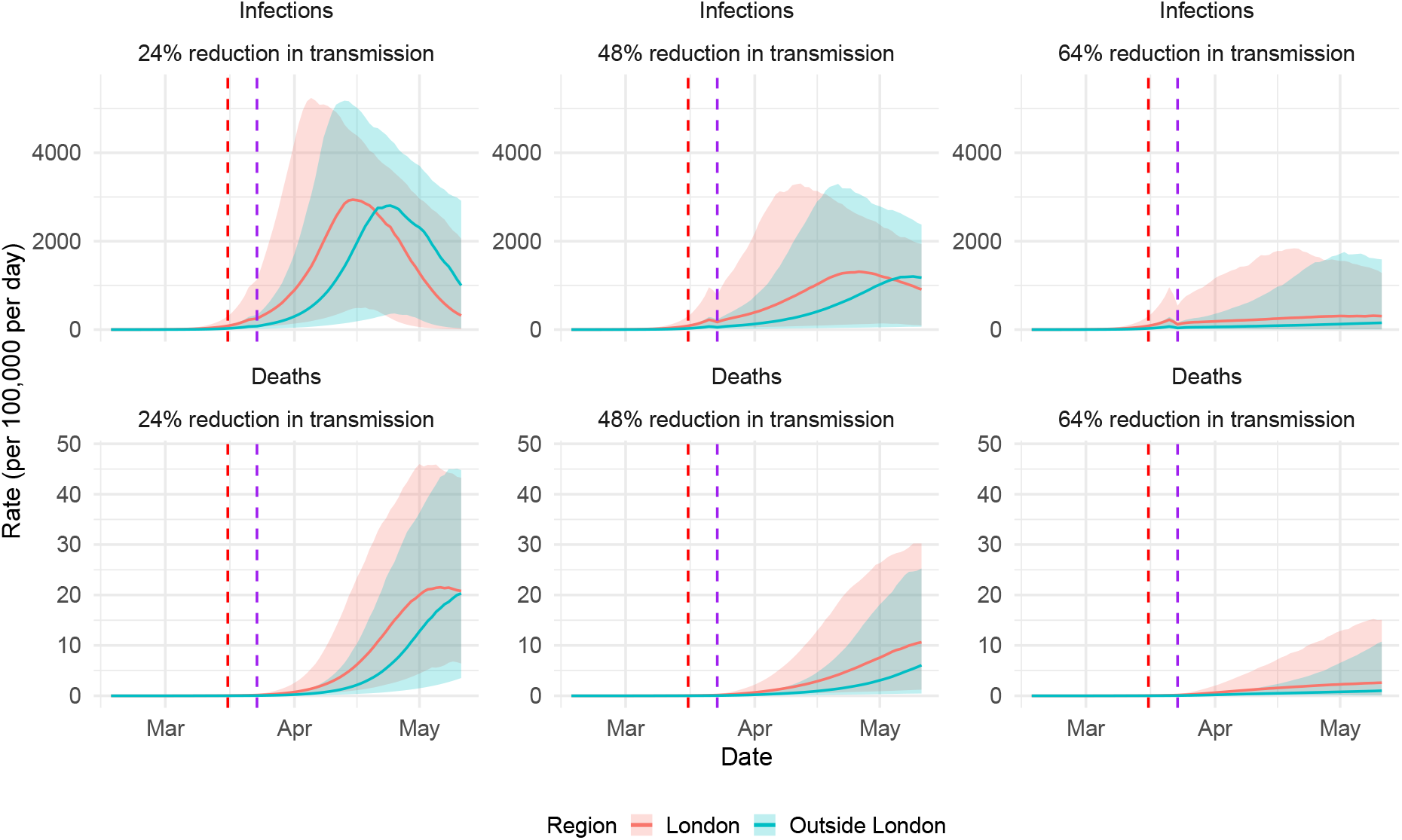
Estimated and projected COVID-19 infections and deaths by efficacy of social restric-tion measures.

The most optimistic scenario in Fig. 2 corresponded to an immediate 64% reduction in transmission. Under this assumption, *R_t_* was estimated to be 1.2 (95% CrI: 0.83–2.1) in London and 1.2 (95% CrI: 0.84–2.2) elsewhere. In this scenario, the probability that the imposed measures were successful in reducing *R_t_* to the threshold of 1 required for declining transmission was only 19% and 17% in the two regions respectively. To be 95% certain that the intervention would lead to a sustained decline in infection, the intervention would need to induce an 81% reduction in transmission. Such a reduction could only be achieved through the implementation of extreme mitigation measures.

### The lock-down period

The number of deaths continued to rise until April 8^th^, particularly in older age-groups (Fig. 1(B)), permitting stratification of the model by both age (8 groups) and region (7 NHS regions). Also new information from serological studies (Fig. 1(C)) started to become available and weekly data downloaded from the Google mobility survey could be used to update contact matrices.

A rhythm for pandemic monitoring was established. The model was run daily, with results feeding into local planning tools as well as the SPI-M consensus view on the state of the pandemic, and with periodic publication of web-reports summarising the latest results (*21*). These outputs included a number of key indicators: regional estimates of *R_t_* and epidemic growth rates r, indicating whether transmission is increasing (*R_t_* > 1) and the rate at which it is increasing (*22*); region and age-specific attack rates (*i.e*. the proportion of the population already infected); and predictions of the burden due to mortality, both in terms of age-specific infection-fatality ratios and number of CoVID19 deaths. Public attention has focused on *R_t_* as a headline figure for the state of the pandemic, but a more complete assessment requires all these indicators. Table 1 presents estimates of a selection of these indicators, giving snapshots of the pandemic state at the three chosen times.

**Table 1:** Table of estimates (with 95% credible intervals attached) for key epidemic parameters and derived quantities.

| Region | 10<sup>th</sup> May |  |  |  |  |  |
| --- | --- | --- | --- | --- | --- | --- |
| | $R_t$ | $r$ | Infections | AR | IFR(overall) | IFR(75+) |
| East | 0.71<br>(0.68–0.74) | -0.07<br>(-0.08– -0.06) | 1,130<br>( 758– 1,660) | 10%<br>(8%–13%) | - | - |
| London | 0.40<br>(0.36–0.43) | -0.18<br>(-0.20– -0.16) | 24<br>(10 – 53) | 20%<br>(16%–26%) | - | - |
| Mids | 0.68<br>(0.65–0.71) | -0.08<br>(-0.08– -0.07) | 1,490<br>(1,080– 2,040) | 11%<br>(9%–15%) | - | - |
| NE&Y | 0.80<br>(0.76–0.83) | -0.05<br>(-0.05– -0.04) | 4,320<br>(3,230– 5,650) | 11%<br>(8%–14%) | - | - |
| North West | 0.73<br>(0.70–0.76) | -0.06<br>(-0.07– -0.06) | 2,380<br>(1,750– 3,160) | 14%<br>(11%–18%) | - | - |
| South East | 0.71<br>(0.68–0.74) | -0.07<br>(-0.08– -0.06) | 1,260<br>(855– 1,810) | 8%<br>(6%–11%) | - | - |
| South West | 0.76<br>(0.72–0.80) | -0.06<br>(-0.07– -0.05) | 739<br>( 438– 1,200) | 5%<br>(4%–6%) | - | - |
| **England** | 0.75<br>0.72–0.77 | -0.06<br>-0.06– -0.05 | 11,400<br>9,150–14,200 | 12%<br>9%–15% | 0.6%<br>0.5%–0.8% | 16%<br>12%–21% |
| Region | 3<sup>rd</sup> June |  |  |  |  |  |
| | $R_t$ | $r$ | Infections | AR | IFR(overall) | IFR(75+) |
| East | 0.94<br>(0.73–1.14) | -0.01<br>(-0.06–0.03) | 1,660<br>( 502– 4,610) | 9%<br>(8%–10%) | - | - |
| London | 0.95<br>(0.72–1.20) | -0.01<br>(-0.07–0.04) | 1,310<br>(247 – 4,670) | 17%<br>(15% – 19%) | - | - |
| Mids | 0.90<br>(0.73–1.07) | -0.02<br>(-0.07–0.01) | 2,460<br>(809– 6,070) | 10%<br>(9%–11%) | - | - |
| NE&Y | 0.89<br>(0.75–1.04) | -0.02<br>(-0.07–0.01) | 2,450<br>(865– 5,870) | 9%<br>(8%–11%) | - | - |
| North West | 1.01<br>(0.83–1.18) | 0.0<br>(-0.04–0.04) | 4,170<br>(1,580– 9,840) | 12%<br>(10%–14%) | - | - |
| South East | 0.97<br>(0.78–1.17) | -0.01<br>(-0.05– -0.03) | 2,420<br>(782– 6,040) | 7%<br>(6%–8%) | - | - |
| South West | 1.00<br>(0.77–1.29) | 0.0<br>(-0.06–0.06) | 778<br>(162– 3,080) | 4%<br>(3%–5%) | - | - |
| **England** | 0.99<br>0.91–1.09 | 0.0<br>-0.02–0.02 | 16,700<br>10,700–25,300 | 10%<br>9%–11% | 0.9%<br>0.8%–1.0% | 23%<br>20%–27% |
| Region | 19<sup>th</sup> June |  |  |  |  |  |
| | $R_t$ | $r$ | Infections | AR | IFR(overall) | IFR(75+) |
| East | 0.80<br>0.60–1.01 | -0.05<br>-0.10–0.00 | 292<br>60–1,050 | 7%<br>6%–8% | - | - |
| London | 0.87<br>0.67–1.12 | -0.03<br>-0.08–0.02 | 837<br>159–3,070 | 17%<br>15%–18% | - | - |
| Mids | 0.82<br>0.64–1.01 | -0.04<br>-0.09–0.00 | 709<br>189–2,040 | 8%<br>7%–9% | - | - |
| NE&Y | 0.76<br>0.58–0.95 | -0.05<br>-0.11– -0.01 | 351<br>84–1,110 | 7%<br>6%–8% | - | - |
| North West | 0.84<br>0.69–1.02 | -0.04<br>-0.08–0.00 | 872<br>255–2,460 | 10%<br>9%–11% | - | - |
| South East | 0.77<br>0.59–0.96 | -0.05<br>-0.10– -0.01 | 342<br>79–1,120 | 6%<br>5%–6% | - | - |
| South West | 0.94<br>0.69–1.21 | -0.01<br>-0.07–0.04 | 312<br>60–1,180 | 3%<br>3%–4% | - | - |
| **England** | 0.88<br>0.79–1.01 | -0.03<br>-0.05–0.00 | 4,260<br>2,370–7,290 | 8%<br>8%–9% | 1.1%<br>0.9–1.4% | 17%<br>14%–22% |

The 10^th^ May section of the table shows the success of the lock-down at curtailing transmission: the *R_t_* in England is now estimated to be 0.75 (95% CrI: 0.72–0.77) having dropped from 2.6 (95% CrI: 2.4–2.9) to 0.61 (95% CrI: 0.57–0.67) at the time of the lock-down, a reduction of 75% (95% CrI: 73%–77%), in line with the anticipation of the pre-lock-down modelling. The ‘growth’ rate for England indicates the daily number of infections were halving every log(2)/r = 11.5 days. London stands out as the region with the highest estimated attack rate (20% of people infected, CrI: 16%–26%), largely due to pre-lock-down levels of infection; the largest reduction in transmission, a drop of 81% (95% CrI: 77%–84%) to *R_t_* = 0.4, 95% CrI: 0.36–0.43, and the steepest rates of decline in both the number of infections (halving every 4 days) and the observed fall in the number of deaths (Fig. 1A). The temporal patterns in infection are disrupted at the lock-down date, with the top row of Fig. 3(A) illustrating the size of this effect for both the London and North West Regions, alongside the estimated *R_t_*.

**Figure 3:**
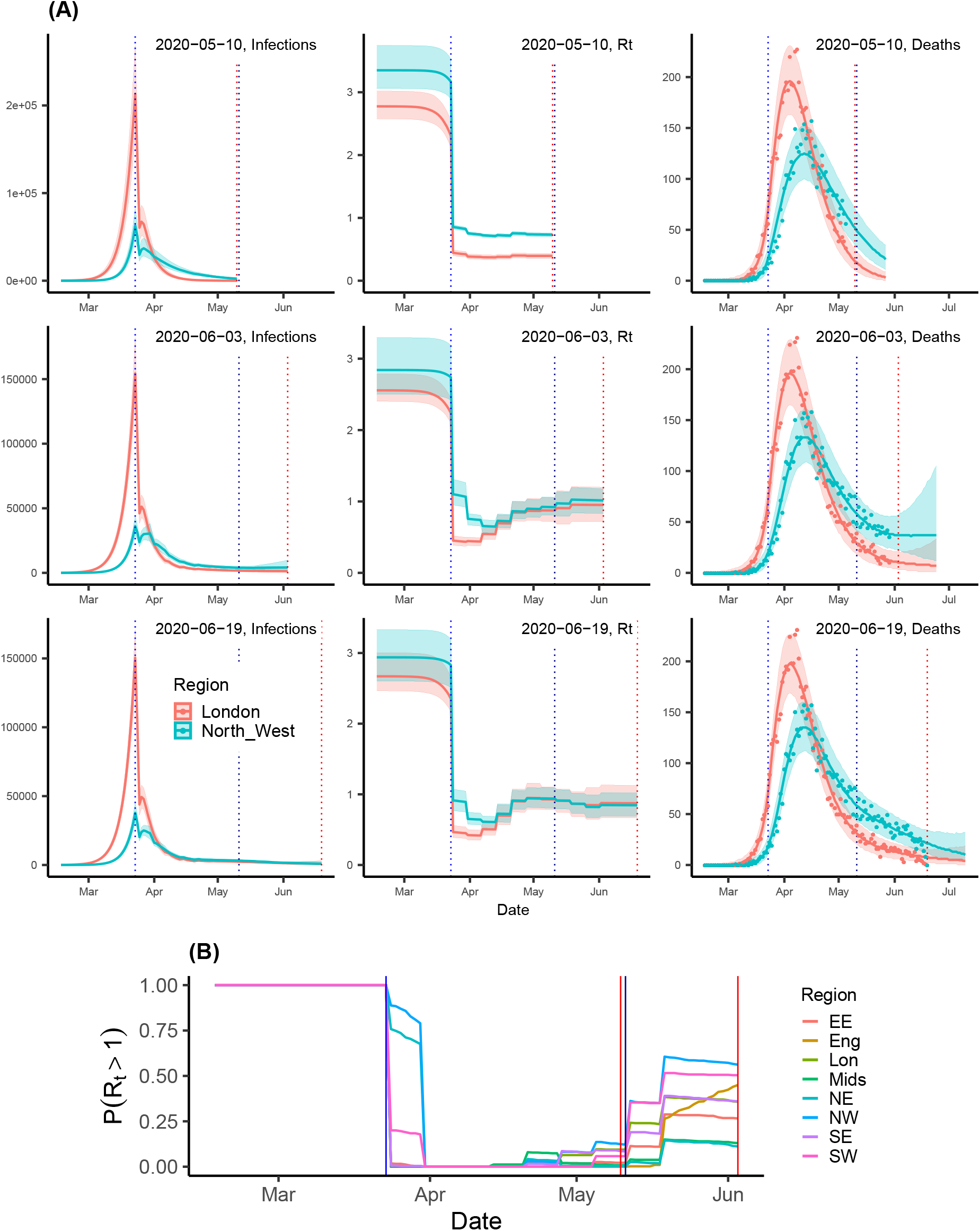
(A) Sequentially obtained estimates infection trajectories; *R_t_*; and three week forecast of the number of deaths by date of analysis (rows). (B) Probability that *R_t_* exceeds 1 as of 3^rd^ June, by region. See SM, Fig. S1 for 10^th^ May8and 19^th^ June. In both plots, vertical blue lines indicate the timings of significant policy changes, vertical red lines indicate the time of analysis.

Launched on the 10^th^ May, the UK COVID 4-level Alert System, co-ordinated by the newly established Joint Biosecurity Centre (JBC) and based on both estimated *R_t_* and current infections, highlighted the key role of these indicators (*7*) in guiding the relaxation of lock-down measures without re-igniting transmission.

### Lock-down Relaxation

The first tranche of relaxations were announced on the 10^th^ May. From this time, accounting for likely changes in behaviour became crucial. In addition, it was clear, from the over-precision of the estimates of both incidence and *R_t_* in the top row of

Fig. 3(A), that the model required a greater flexibility to capture such changing behaviours. From an appropriately adapted model (see SM ‘Transmission model’ for details), we estimated that at the 3^rd^ June, *R_t_* for England reached 0.99 (95% CrI: 0.91–1.09), with the probability of exceeding the value of 1 rapidly increasing over time (see Fig. 3(B)). This figure masks regional heterogeneity in transmission. The North West and the South West were characterised by *R_t_* values above 1 (with a probability *R_t_* > 1 of over 50%, Fig. 3(B)) and growth rate estimates encompassing positive values. For the North West, we estimated 4,170 (95% CrI:1,580–9,840) daily infections, the highest number in the country (see Table 1). The step changes in the plots of *R_t_* over time in Fig. 3 for 10^th^ May are entirely due to changes in the Google mobility matrices. Looking at the equivalent plot for the 3^rd^ June analysis, over the same interval, the step changes are larger. This difference in the level of fluctuation over time suggests that the increases in *R_t_* are too large to be solely attributed to mobility-driven changes in the contact matrices. Furthermore, for the North West, the drop in *R_t_* around the lock-down is not as sharp as in London, but rather staggered over three weeks. This might suggest a different response to the lock-down in these two regions, which we had previously not been able to identify. The estimated steady resurgence of *R_t_* in the North West ultimately led to a policy change, delaying the staged re-opening of schools (*23*).

Continuing to monitor the pandemic evolution in the post-lock-down era, we adapted the model to incorporate new evidence on differential susceptibility to infection by age (*24*) (see SM, around equation (7)). Results from 19^th^ June data (Table 1, Fig. 3(A)) show lower estimates for *R_t_*, negative growth rates and the estimated number of infections in England decreasing to 4,300 (95% CrI: 2,400 - 7,300). There is still regional heterogeneity, with two regions for which the credible intervals for *R_t_* exclude 1 (North East & Yorkshire, and the South East); and the probability that the epidemic is growing is 30% in the South West and below 15% in each of the other regions (Fig S1(B), SM). Throughout we have been estimating age-specific infection-fatality ratios (see Tables 1 and S4). Allowing for differential age susceptibility, the age-specific estimates of the infection-fatality ratio fall to 17% (95% CrI: 14%–22%) in the over-75s (from 23%, 95% CrI: 20%–27%) with a rise to 2.9% in the 65–74ys, (see Table S4) and to 1.1% (95% CrI: 0.9%–1.4%) from 0.9% (95% CrI: 0.8%–1.0%) overall. These less severe estimates (in comparison to the 3^rd^ June analysis) led to the UK Chief Medical Officers agreeing with a JBC recommendation that the alert level should be downgraded to level three.

## Discussion

Since the 19^th^ of June, weekly updates have continued to be posted online (*21*). Recent estimates of the IFR have varied in the region of 0.9% to 1.4% overall and 14% to 19% in the over-75s. *R_t_* is approaching the value of 1 in most regions without exceeding it, which, together with a decreasing number of daily infections indicates an epidemic still in decline, although local outbreaks are being increasingly detected (*25*). The estimates presented here are consistent with the SPI-M consensus on the values of *R_t_* both nationally and regionally (*26*). Incidence estimates can be contrasted with estimates from community cross-sectional studies. In a report of the 9^th^ July, the Office for National Statistics (ONS) estimates 1,700 (range 700–3,700) new daily infections over the two weeks leading up to 4^th^ July (*27*), while the ZOE study (*28*) reports an average 1,470 infections per day over a similar period. In our work incidence ranges from a central estimate of 4,200 daily infections down to 3,500 over the period. These are not incongruous to the ZOE app estimates, which only records symptomatic infection and require some scaling to derive an estimate for total infections. The ONS estimate is likely to be an under-estimate as the survey does not include individuals in institutionalised settings e.g. care homes, where incidence may be far higher than in the community. The degree to which this is an under-estimate, however, is unclear.

The ONS also reports (*29*) that 6.3% (95% CrI: 4.7%–8.1%) of individuals showed the presence of antibodies to the COVID-19 in blood sera samples (as of 19^th^ June), a little lower than the 8% estimated attack rate for England. This discrepancy may well be due to the timing of the samples, due to the waning of the antibody response over time. Estimates of attack rates (see Table 1) show that our belief on the proportion of the population that has been infected is being revised downwards at each sequential analysis. This, together with the emerging evidence of waning immunity (*30*), paint a muddied picture in term of potential for a population-level resurgence in infection.

On the 4^th^ July there was a significant step change in the gradual relaxation of pandemic mitigation measures as leisure facilities, tourist attractions, pubs and cafes all became accessible to the public once again (*31*). The impact of this is still largely to be observed and assessed. If the concerning worse-case scenario presented for the Winter in the recent publication of the Academy of Medical Science (*32*) comes through, we will be facing a very challenging second wave. Our monitoring tool, continuously adapted to incorporate the accumulating surveillance data, will be key to providing quantification of all aspects of any resurgence.

## Data Availability

The data used in the manuscript are routinely collected surveillance data on the ongoing CoVID-19 pandemic. We are in the process of agreeing and appropriate level of aggregation under which these data can be made available in a public repository.

## Acknowledgements

This work was supported by the Medical Research Council (Unit programme number MC UU 00002/11) in partnership with Public Health England. Prior to the pandemic, the project was developed under a grant from the National Institute for Health Research (HTA Project: 11/46/03). We gratefully acknowledge the access to the data from the United Kingdom Time Use Survey through the UK Data Service (http://doi.org/10.5255/UKDA-SN-8128-1). We ac-knowledge the support of the PHE Epidemiology Cell in consistently providing the data streams used. We would also like to thank Dr Nigel Gay for the initial development of the model and constructive comments on the manuscript. This work has also benefited from insightful discussions with Dr Petra Klepac.

The core membership of the PHE Modelling Cell consists of: Abbygail Jaccard, Andre Charlett, Anna Rance, Anne Presanis, Archana Purohit, Brian Ferguson, Brodie Walker, David Mustard, Declan Bays, Dianne Addei, Emilia Vynnycky, Emily Agnew, Emma Bennett, Emma Gillingham, Hannah Williams, Ian Hall, James Lewis, Jonathan Carruthers, Joseph Shingleton, Joshua Blake, Judith Field, Martin Grunnill, Matt Edmunds, Matt Hennessey, Nick Gent, Peter White, Simona Baracaia, Stephanie Shadwell, Steven Dyke, Thomas Finnie, Virginia Cox and Xu-Sheng Zhang.

## Supplementary Materials

### Methods

#### Surveillance data

The UK confirmed its first case of CoVID infection, imported from China, on the 30^th^ January, with the first CoVID-linked death reported on the 6^th^ March. Initially deaths were monitored by date of report, (*33*) with a full line-listing of deaths being made available by PHE from the 21^st^ March, enabling tracking of the pandemic using actual dates of death.

Prior to this time, the most complete data were the time series of reported cases, individuals who have a returned a PCR-positive swab sample. (*33*) Initial analyses of the pandemic were based on either or both of these datasets, attempting to quantify the emerging pandemic but also to monitor the degree of coherence between the datasets. However, rates of testing and the country’s testing capacity were significantly increasing, (*34*), whilst government policy was changing the testing threshold. These led to a significant ascertainment bias to the confirmed case data which in turn would lead to estimated incidence curves based on these data to be significantly skewed.

A third data source was the testing of residual blood sera from blood donors in England via National Health Service Blood and Transplant (NHSBT). These data inform the population prevalence of antibodies, informing the fraction of the population that have ever been infected. From repeated follow-up of known infections, 74.7% (*35*) of infections return a positive result for the presence of antibodies within 21 days of the onset of symptoms

#### Transmission Model

An age stratified transmission model is fitted to each region under study simultaneously with the sharing of some global parameters. Within each region, the transmission dynamics are governed by a system of ordinary differential equations, discretised to give the following set of first order difference equations:

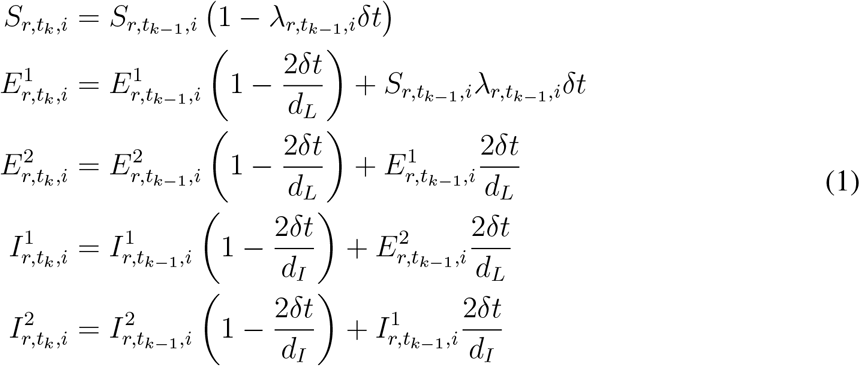

where: 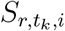, 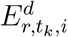, 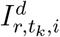, *d* =1, 2 represent the time *t_k_,k* = 1,…, *K*, partitioning of the population of individuals in a region *r,r* = *1, …, n_r_*, in age-group *i,i* = *1,…, n_A_*, into *S* (susceptible), *E* (exposed) and *I* (infectious) disease states. The mean latent and infectious periods are *d_L_* and *d_I_* respectively; and 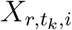 is the time- and age-varying rate with which susceptible individuals become infected. Time steps of *δt* = 0.5 days are chosen to be sufficiently small relative to the anticipated latent and infectious periods. New infections are generated as

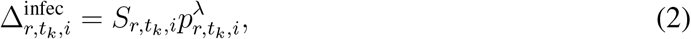

where

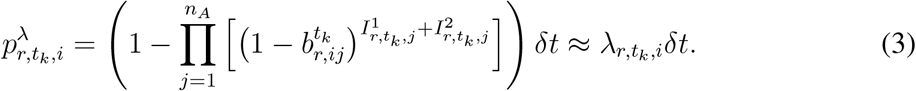

Here, 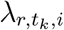 is the probability of a susceptible individual in region *r* of age group *i* being infected by an infectious individual in age group *j* at time *t_k_*. It is a function of:

- a set of time-varying contact matrices 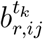, the entries of which describe the expected number of contacts between any two individuals of the different strata within a single time unit.
- 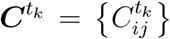, a region-specific matrix, the (*i, j*)^th^ element of which gives the relative susceptibility of someone in age-group i to an infection from an infectious individual in age-group j assuming contact between the two.
- *β_tk_,_r_*, a time-varying parameter encapsulating further temporal fluctuation in transmission that applies to all ages.
- *R*_0_,*_r_* are the initial reproduction numbers for the pandemic in each region at time *t*_0_
- 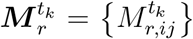 are the dominant eigenvalues of the initial next-generation matrices, Λ_0_,*_r_* such that

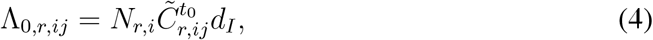

where *N_r_,_i_* is the population size in region r and age-group i, and *cr^k^* are a set of matrices defined by

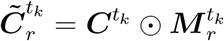

where the ⊙ notation indicates element-wise multiplication, s.t *A* = *B* ⊙ *C* if *A_ij_* = *B_ij_C_ij_*.

As described in the following sub-section, the *C^tk^* matrices encode the information contained about contact rates between different age groups derived from the POLYMOD study (*18*), Google mobility and the time-use survey. The 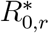 matrices capture any misspecification of these matrices in terms of the changing pattern of infection between the age groups, whereas the 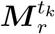 parameters account for misspecification of the changing scale of transmission over time as described by the matrices.

In the main manuscript, a number of model developments are identified that were implemented in between the analyses presented. The general expression of 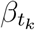 is:

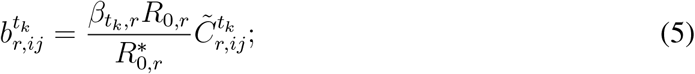

but each of the models implemented constitute some simplification of this expression. These model developments are itemised here, starting with the earliest, and most simple, analysis:

- **20^th^ March:** No age dimension to the analysis, so transmission was simply broken up with a changepoint at *t_k_* = *t_lock_*. The dynamics of the two regions (London, Outside London) differed only in that they had a differing initial seeding of infection. Here 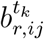 and 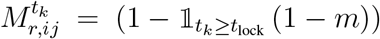, giving m the interpretation as a global change in the susceptibility to infection given contact with an infectious individual. From (*4*), 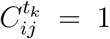. Simplifying (*5*):

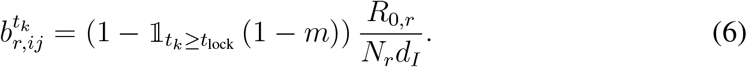
- **10^th^ May**: Analysis is now age-specific. Reproduction number *R*_0_*,_r_* and susceptibility parameter *m_r_* now region-specific. The matrices 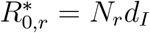 are a matrix extension of the 20^th^ March analysis:

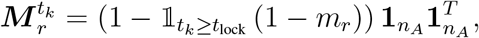

where the susceptibility parameter *m_r_* now has region dependence, 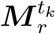 is a vector of ones of length *n_A_*, and 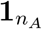 is a corresponding matrix of ones. This has no age dependence, and so it is assumed that the POLYMOD and Google-derived matrices, 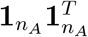, accurately account for changing patterns of infection over time). Again (*4*) can be simplified to:

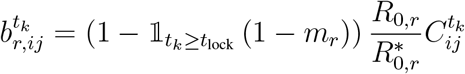
- **3^rd^ June:** Introduction of the time-varying transmissibility parameter, 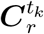 It was decided to change these piecewise with weekly changepoints. Denote *w_k_* ≡ *w*(*t_k_*) giving the week in which time *t_k_* falls. Over time these are not allowed to vary unconstrained and a smoothing is imposed by assuming, *a priori* that they develop according to a random-walk.

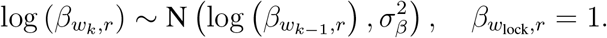 Where 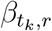 is the rate that applies in all weeks up to and including the first week of the lock-down. As all other components of the model remain unchanged, the form for 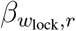 is:

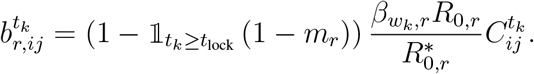
- **19^th^ June**: Separate levels of susceptibility are introduced for the over-75s, both before and after the lock-down to account for some lack of fit. To characterise this, we need to define two matrices, 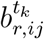 and 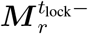 that apply before and after *t_lock_* respectively,

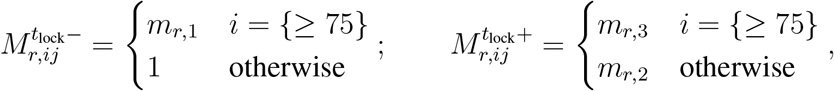

such that>

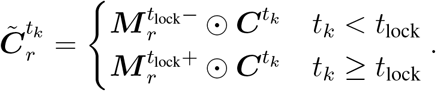 We would then feed back into equation (*3*):

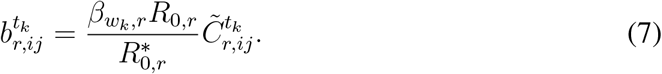

#### Calculating *R_t_*

We estimate a region-specific parameter describing the initial exponential growth in infections, denoted *ψ_r_*. Using the formula of Wearing et al. (2005), this is related to the initial reproduction number *R*_0,_ *_r_* via the formula

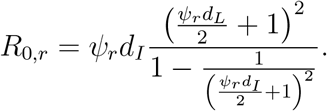

Over time the value of the reproduction number will change as contact patterns shift and the supply of susceptible individuals deplete. The formula for the time-*t* reproduction number is

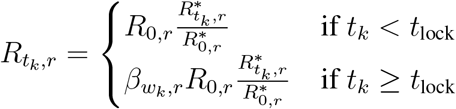

where 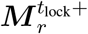 is the dominant eigenvalue of the time *t_k_* next-generation matrix, Λ*_k_,_r_*, with elements:

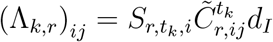

To get an ‘all England’ value for 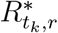 a weighted average of the regional 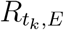 is calculated, where the weights are given by the sum of the infections in each region:

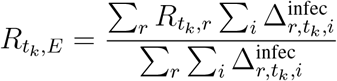

### Contact matrices

Time dependent contact matrices are based on location-specific POLYMOD matrices (where locations include “at work”, “at home”, “on transport” etc), combined with the time-use survey (*19*) to identify 18 different activities, including school, work, social visits, shopping etc.. The traditional POLYMOD matrices are used until 23^rd^ March, the time of the lock-down (*18, 36*). From this point on, the Google mobility and time-use survey data were used to calculate proportionate reductions in the location-specific POLYMOD matrices, which are summed together to give a weekly-varying contact matrix, 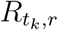.

Following (*19*) we identified 18 different activities (Table S1). Some of these activities were not allowed during lock down and, therefore, assumed to have been stopped. For other activities we used the most relevant data source. For some activities no suitable data source was available. In that case we used the retail and recreation mobility data provided by Google, because this data was assumed to best represent the general adherence level in the UK. For example, visits are unlikely to have stopped completely. Instead the retail and recreation level is used (Table S1). Note that the activities which in a well mixed model have the most effect on the base reproductive number are school, work, visits and unspecified (*19*).

**Table S1:** The data sources used for the different activities identified by the time use survey. Values represent the assumed activity level. Where direct data was available we used other (live) data sources. Here GM represents the relevant Google mobility category.

| activity | Data source |
| --- | --- |
| alone | 1 |
| bars and restaurants | 0 |
| bed | 1 |
| cultural | 0 |
| exercise indoors | 0 |
| home | 1 |
| library | 0 |
| school | Attendance records |
| visit | GM Retail & recreation |
| parks | GM Parks |
| holiday | GM Retail & recreation |
| shopping | GM Retail & recreation |
| exercise outdoors | GM Retail & recreation |
| exercise unspecified | GM Retail & recreation |
| shopping essential | GM Grocery & pharmacy |
| unspecified | GM Retail & recreation |
| work | GM Workplace |
| transport | GM Transit stations |

**Table S2:** The fraction of contacts at home attributed to members of the household.

| AgeGroup | Contacts at home | Max contacts with household members | fraction |
| --- | --- | --- | --- |
| [0, 1) | 5.643 | 2.857 | 0.506 |
| [1, 5) | 4.295 | 2.808 | 0.654 |
| [5, 15) | 4.980 | 2.970 | 0.596 |
| [15, 25) | 4.206 | 2.675 | 0.636 |
| [25, 45) | 4.125 | 2.388 | 0.579 |
| [45, 65) | 3.767 | 1.524 | 0.405 |
| [65, 75) | 3.614 | 1.045 | 0.289 |
| [75, +) | 3.571 | 0.714 | 0.200 |

### School attendance

School size and age range is publicly available for England. We used this to calculate the number of students for each school in the modelled age groups, based on the assumption that the students were evenly distributed across different school years. We also had access to attendance levels over time for some schools. This data was then used to calculate the average weekly attendance level in each local authority, weighted by the size of the school in that age group. Finally, we combined these values to calculate attendance in England, weighted by local authority population size. Not all schools reported attendance every week, as a result the attendance by local authority was, in rare cases, only based on the report of 1 or more small schools. To ensure that these did not skew our results, we ignored any attendance estimate based on schools with less than 100 students in total.

### Visits at home

(*19*) used the time usage data for visits at home to estimate the number of visit related contacts at home. We found that this underestimates the number of visitors and instead used the POLYMOD data on household size to estimate the fraction of contacts at home with other household members versus the contacts with others (e.g. visitors). First, we extracted the mean number of contacts at home (Table S2). Next, we limited numbers of contacts at home (*c_i_*) to household size (*h_i_*) minus one 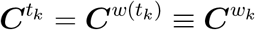, i.e. the maximum number of contacts any participant (*i*) can have with just household members and calculated the mean contacts based on that value. Note that this provides a conservative estimate of the fraction of contacts from visits, because some participants will not have met all their household members during the day.

### Google mobility data

During the pandemic Google provided aggregated mobility data from Android phones for many countries (https://www.google.com/covid19/mobility/). The mobility data gives an indication of the activity level for 5 different activities: retail and recreation, grocery and pharmacy, parks, transit stations, workplaces and residential. The UK data was further subdivided into activity by local authority, by matching the Google provided locations to local authority districts in England. This data was then combined, weighted by population size. Finally, the daily values were averaged by week, to produce weekly activity levels.

## Likelihood

Counts of deaths are assumed to have a negative binomial distribution. If 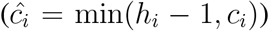 is the number of observed deaths on day *t_k_* in age group *i* in region *r*, then the expected number of deaths are derived using (*2*), an assumed-known distribution of the time from infection to death from COVID-19, *f* and an estimated age-specific infection-fatality ratio *p_i_*:

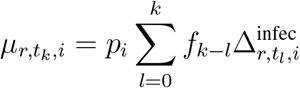

where *f_l_* gives the probability the death occurs on the *l*^th^ day after infection. Then we assume

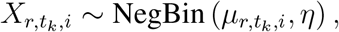

where *η* is a dispersion parameter such that 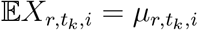 and 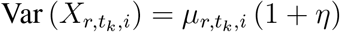.

The serological data is dependent on two parameters, the sensitivity and the specificity of the serological testing process, *k_sens_* and *k_spec_* respectively. If, on day *t_k_*, 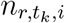 blood samples are taken from individuals in region r and age-group i, and the observed number of positive tests is 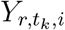, then

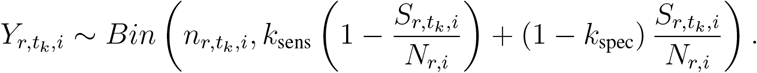

### Inference

Table S3 contains a full list of parameters. The top portion refers to the unknown parameters that are estimated and the bottom portion contain parameters that have been fixed. Wherever possible, a brief justification of the chosen prior/value is given.

**Table S3:** Model parameters with assumed prior distributions or fixed values Name Prior source

| Name | Prior source |
| --- | --- |
| Over-dispersion, $\eta$ | Uninformative $\Gamma(1, 0.2)$ |
| Mean infectious period, $d_I$ | $2 + \Gamma(1.43, 0.549)$, based on Li et al (2020). |
| All contact matrix multipliers, $m_{r,l}, l = 1, \dots, 3$ | Uninformative $\Gamma(4, 4)$ |
| Exponential growth, $\psi_r$ | $\Gamma(31.36, 224)$, through matching to an uninformative $R_0$ given sampled values of $d_I$ and assumed value of $d_L$. |
| Initial infection, $I_0$ | Uninformative. To be described. |
| Infection-fatality rate $p_i$ | Based on Verity et al. (2020). |
| Serological test sensitivity, $k_{\text{sens}}$ | Based on convalescent sera, $\beta(71.5, 29.5)$. |
| Serological test specificity, $k_{\text{spec}}$ | Based on pre-COVID sera, $\beta(777.5, 9.5)$. |
| Step-size on log-scale of weekly variation in transmission, $\sigma_\beta$ | Informative $\Gamma(1, 100)$. |
| Mean latent period, $d_L$ | 4 days. |
| Mean (s.d) incubation period | 4.0 (1.41) days. |
| Mean (s.d) time to death from symptom onset | 15.0 (12.1) days. |

Estimation is conducted in a Bayesian framework by combining the prior information and likelihoods specified above, to derive posterior distributions for the unknown parameters and any functions of such parameters. Sampling from these posterior distribution is carried out through Markov chain Monte Carlo (*13,14*). The most recent analysis featured in this paper was based on 900,000 iterations, with an initial adaptive phase of 45,000 iterations within a burn-in period of 90,000 iterations. Parameter estimates are based on the full sample following burn-in thinned to retain every 125 iterations, and projections are based on a sample thinned to every 250 iterations. All central estimates are pointwise medians of quantities calculated on the basis of this sample, and uncertainty is expressed through 95% credible intervals (CrI) derived from the 2.5% and 97.5% quantiles.

## Additional Results

In Fig. S1, we present the region-specific probabilities that *R_t_* > 1. In the 10^th^ May analysis, the plot is binary - *R_t_* is definitely above the threshold before the lockdown, and we estimated the reduction in transmission to be sufficient to reduce it to below the threshold with near certainty. However, by the time of the 19^th^ June analysis, there is much greater flexibility and uncertainty in the evolution of *R_t_* over time and correspondingly, after 3–4 weeks beyond the lock-down, *R_t_* can no longer be said with certainty to be below 1. From this time there is an initial resurgence in the probability *R_t_* > 1 which then plateaus from mid-May onwards. By the end of the period, there is the possibility that *R_t_* > 1 in both the South West and in London, but it remains unlikely.

Table S4 gives the full estimation of the infection-fatality ratios only partially presented in Table 1.

Fig. S2 shows the goodness-of-fit to the death data by age, for each of the three post-lockdown analyses considered. In the 3^rd^ June analysis, there is a consistent under-estimation of the number of deaths from the beginning of May in the over-75s, with corresponding over-estimation in the younger age-groups. The model adaptation to allow for a different susceptibility to infection for the over-75s appears to rectify this lack of fit.

**Figure S1:**
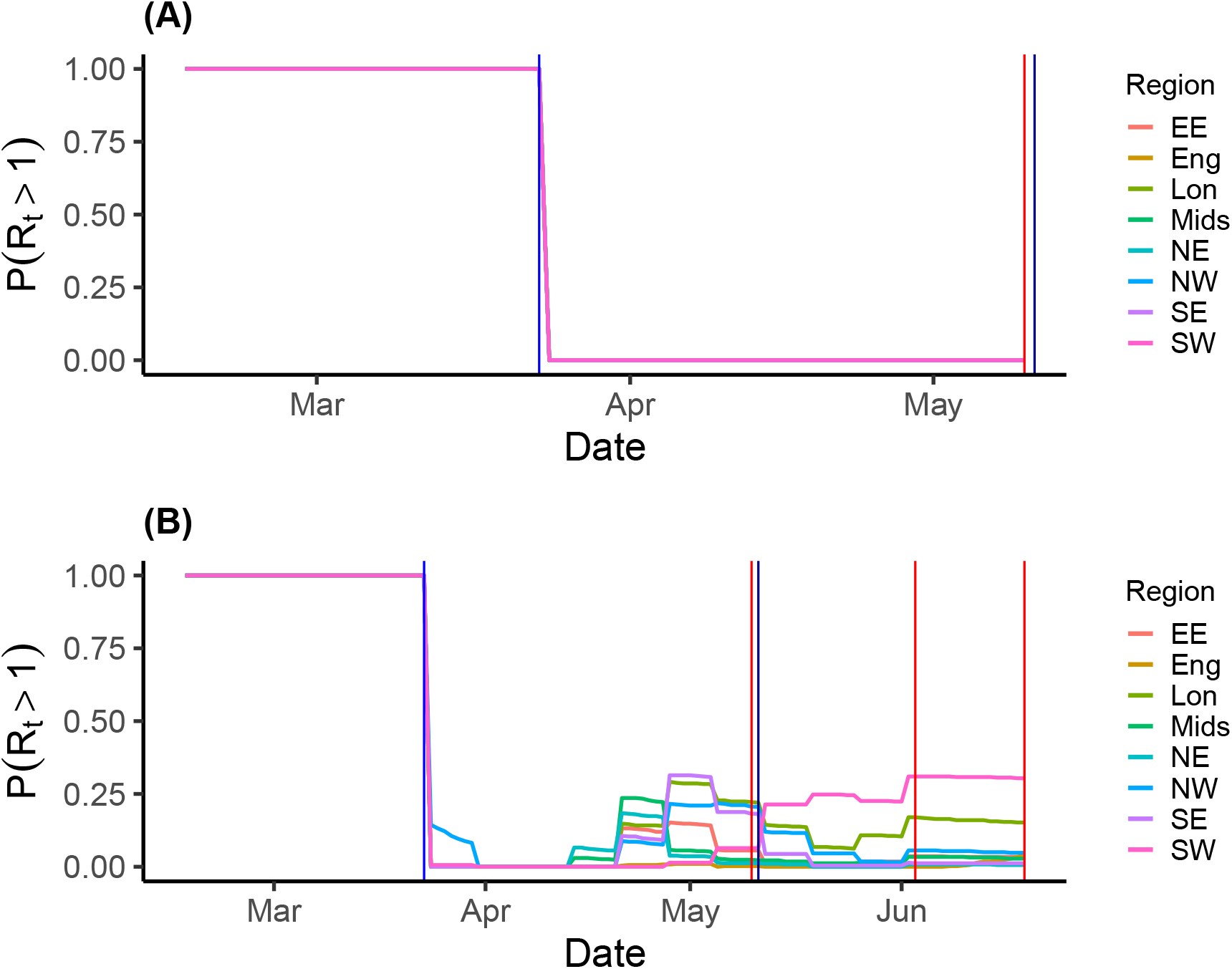
Equivalent plots to Fig. 3(B) for the analysis on (A) 10^th^ May, and (B) 19^th^ June.

**Table S4:** Estimates for the age-specific fatality ratio as they change over time

| Age group (yrs) | 10<sup>th</sup> May | 3<sup>rd</sup> June | 19<sup>th</sup> June |
| --- | --- | --- | --- |
| < 5 | 0.0005%<br>(0.00009%–0.0017%) | 0.0004%<br>(0.00002%–0.0016%) | 0.0005%<br>(0.00008%–0.002%) |
| 5–14 | 0.0006%<br>(0.0002%–0.0013%) | 0.0010%<br>(0.0005%–0.0018%) | 0.0013%<br>(0.0007%–0.0023%) |
| 15–24 | 0.0032%<br>(0.0019%–0.0049%) | 0.0039%<br>(0.0026%–0.0057%) | 0.0043%<br>(0.0029%–0.0062%) |
| 25–44 | 0.018%<br>(0.013%–0.024%) | 0.024%<br>(0.020%–0.029%) | 0.029%<br>(0.025%–0.034%) |
| 45–64 | 0.28%<br>(0.21%–0.37%) | 0.36%<br>(0.32%–0.42%) | 0.44%<br>(0.40%–0.49%) |
| 65–74 | 1.8%<br>(1.3%–2.3%) | 2.3%<br>(2.0%–2.7%) | 2.9%<br>(2.6%–3.2%) |
| > 74 | 16%<br>(12%–21%) | 23%<br>(20%–27%) | 17%<br>(14%–22%) |
| **Overall** | 0.63%<br>(0.49%–0.81%) | 0.88%<br>(0.77%–1.0%) | 17%<br>(0.90%–1.4%) |

**Figure S2:**
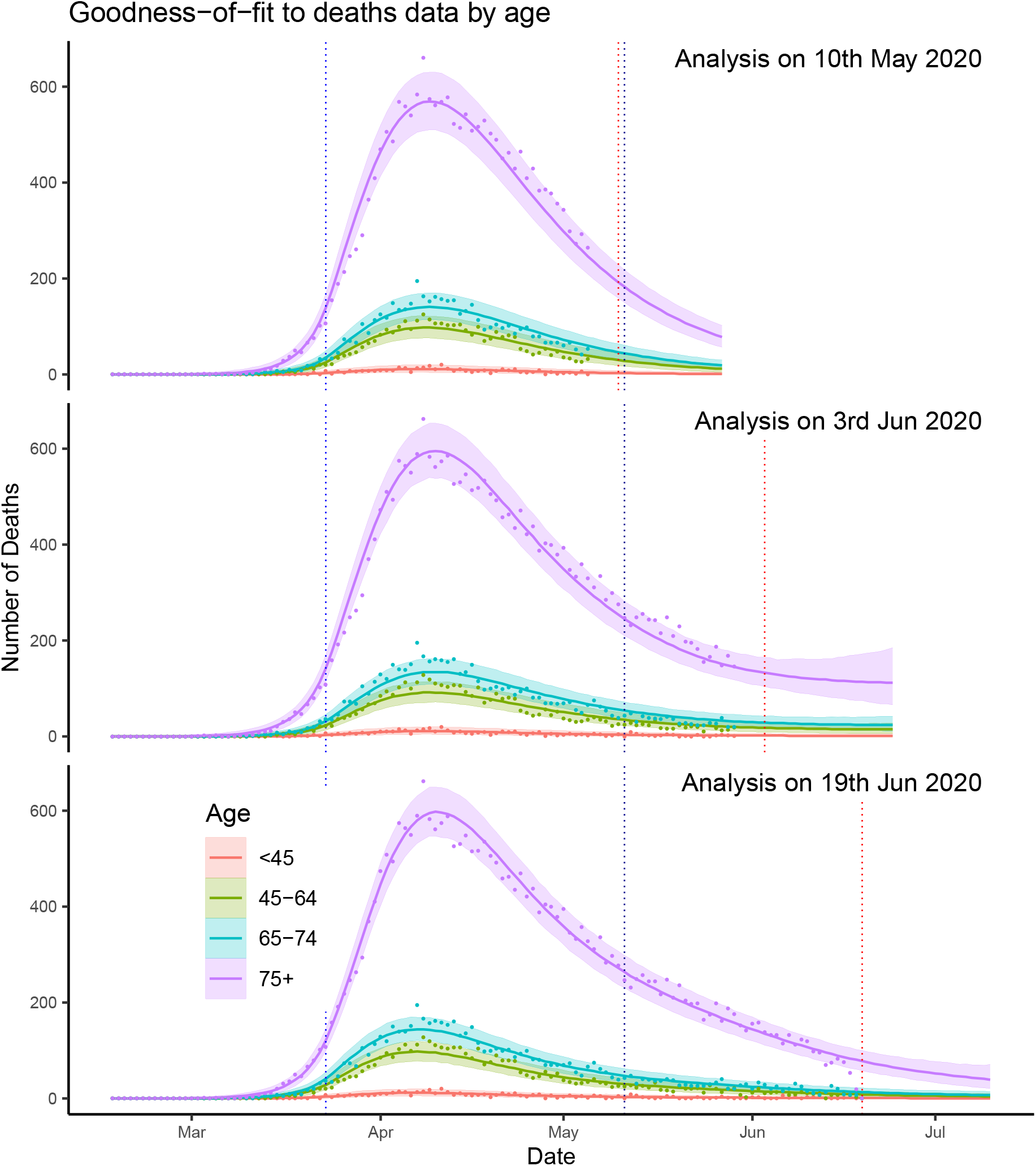
Goodness-of-fit to the deaths data by age on the 10^th^ May, 3^th^ June and 19^th^ June

